# Searching for Dermatology Information Online using Images vs Text: a Randomized Study

**DOI:** 10.1101/2024.10.25.24316155

**Authors:** Justin D Krogue, Rory Sayres, Jay Hartford, Amit Talreja, Pinal Bavishi, Natalie Salaets, Kimberley Raiford, Jay Nayar, Rajan Patel, Yossi Matias, Greg S Corrado, Dounia Berrada, Harsh Kharbanda, Lou Wang, Dale R Webster, Quang Duong, Peggy Bui, Yun Liu

## Abstract

**Background:** Skin conditions are extremely common worldwide, and are an important cause of both anxiety and morbidity. Since the advent of the internet, individuals have used text-based search (eg, “red rash on arm”) to learn more about concerns on their skin, but this process is often hindered by the inability to accurately describe the lesion’s morphology. In the study, we surveyed respondents’ experiences with an image-based search, compared to the traditional text-based search experience.

**Methods:** An internet-based survey was conducted to evaluate the experience of text-based vs image-based search for skin conditions. We recruited respondents from an existing cohort of volunteers in a commercial survey panel; survey respondents that met inclusion/exclusion criteria, including willingness to take photos of a visible concern on their body, were enrolled. Respondents were asked to use the Google mobile app to conduct both regular text-based search (Google Search) and image-based search (Google Lens) for their concern, with the order of text vs. image search randomized. Satisfaction for each search experience along six different dimensions were recorded and compared, and respondents’ preferences for the different search types along these same six dimensions were recorded.

**Results:** 372 respondents were enrolled in the study, with 44% self-identifying as women, 86% as White and 41% over age 45. The rate of respondents who were at least moderately familiar with searching for skin conditions using text-based search versus image-based search were 81.5% and 63.5%, respectively. After using both search modalities, respondents were highly satisfied with both image-based and text-based search, with >90% at least somewhat satisfied in each dimension and no significant differences seen between text-based and image-based search when examining the responses on an absolute scale per search modality. When asked to directly rate their preferences in a comparative way, survey respondents preferred image-based search over text-based search in 5 out of 6 dimensions, with an absolute 9.9% more preferring image-based search over text-based search overall (p=0.004). 82.5% (95% CI 78.2 - 86.3) reported a preference to leverage image-based search (alone or in combination with text-based search) in future searches. Of those who would prefer to use a combination of both, 64% indicated they would like to start with image-based search, indicating that image-based search may be the preferred entry point for skin-related searches.

**Conclusion:** Despite being less familiar with image-based search upon study inception, survey respondents generally preferred image-based search to text-based search and overwhelmingly wanted to include this in future searches. These results suggest the potential for image-based search to play a key role in people searching for information regarding skin concerns.

## Introduction

Skin conditions affect a large proportion of the population, causing significant anxiety and morbidity worldwide^1,2^. Early identification of the condition is an important step for effective treatment and management^3^. Unfortunately, access to physicians for these conditions is often limited, especially in areas where dermatologists may be lacking^4,5^; it has been reported that more than 3 billion people in developing countries lack adequate access to dermatology care^6^. Individuals are then often left to themselves to gather information and determine their next steps. While the internet contains a large amount of information about various skin conditions, finding the most relevant information is traditionally done via text-based search. Unfortunately, without relevant training, it is difficult for users to learn the relevant terminology and patterns necessary to describe their skin-based lesion precisely (e.g., pustules, macules)^7,8^, therefore reducing the precision of the search results.

Recent advances in computer vision have enabled a new type of search experience, in which users submit images as queries and receive relevant matching results^9^. Additionally, previous work has shown that it is possible for machine-learning based systems to accurately identify skin conditions from user-submitted photographs^10–14^. Such a system of image-based search tuned for skin conditions in combination with the increasing prevalence of smartphone usage worldwide could enable billions more to access appropriate sources of information regarding myriad skin conditions. To this end we have updated an existing image search application (Google Lens) to allow users to submit a photo of a skin-based condition and receive an image-grid of dermatology conditions with similar appearances sourced from the world wide web. Users are then free to examine these images, explore the ones that most resemble their issue, and conduct further searches as desired via traditional text-based search.

In this study we examine respondents’ experience with both this image-based system and traditional text-based search. Satisfaction and preferences after each experience is recorded and compared. To examine if certain subgroups feel differently about the feature, we further examine satisfaction and preferences by self-reported demographic subgroups, comparing the results amongst different age groups, genders, ethnicities, skin tones, etc. to the general population of respondents. We hypothesize that respondents will prefer image-based search to text-based search and, having tried it, will want to include it in future searches for skin conditions.

## Methods

### Recruitment and study design

This survey-based research study’s protocol was reviewed and deemed exempt from further oversight by Advarra institutional review board (IRB). Individuals were consented and enrolled from the Qualtrics panel and then randomized into either text-based search (Google Search) first or image-based search (Google Lens) first, both via the Google mobile app. In other words, all respondents used both text-based search and image-based search, but randomized orders of which modality they tested first. Recruitment had an initial stratification to enrich for older age, balance the genders, and diversify based on race/ethnicity (using US census breakdowns as targets). These constraints were relaxed after enrollment slowed, to achieve the target enrollment. Exclusion criteria included those outside of the US, those who did not speak English, and those lacking a visible concern that they were willing to do a search of.

Prior to performing the image-based and text-based searches, self-reported basic demographic information was collected, including gender, race, ethnicity, age bracket, and Fitzpatrick skin type. Respondents were also asked to rate their tech literacy and educational levels. Additionally, respondents were asked a few basic details about their concern, including how long they had noticed it, and whether or not they had a previous diagnosis. Respondents were also asked to rate how familiar they were with image-based search and text-based search.

After performing their search, respondents were asked to submit a screenshot of their search results to ensure that they had performed the correct type of search. Successful completion of the two types of searches were verified by Qualtrics using the screenshots, and only those who completed both types of searches were included in the study. Figure 1 shows a STARD diagram of flow of study respondents. No identifying data were collected in this study; respondents were instructed not to submit identifying or sensitive information, and data were further scrubbed for such information by Qualtrics prior to transfer.

**Figure 1:**
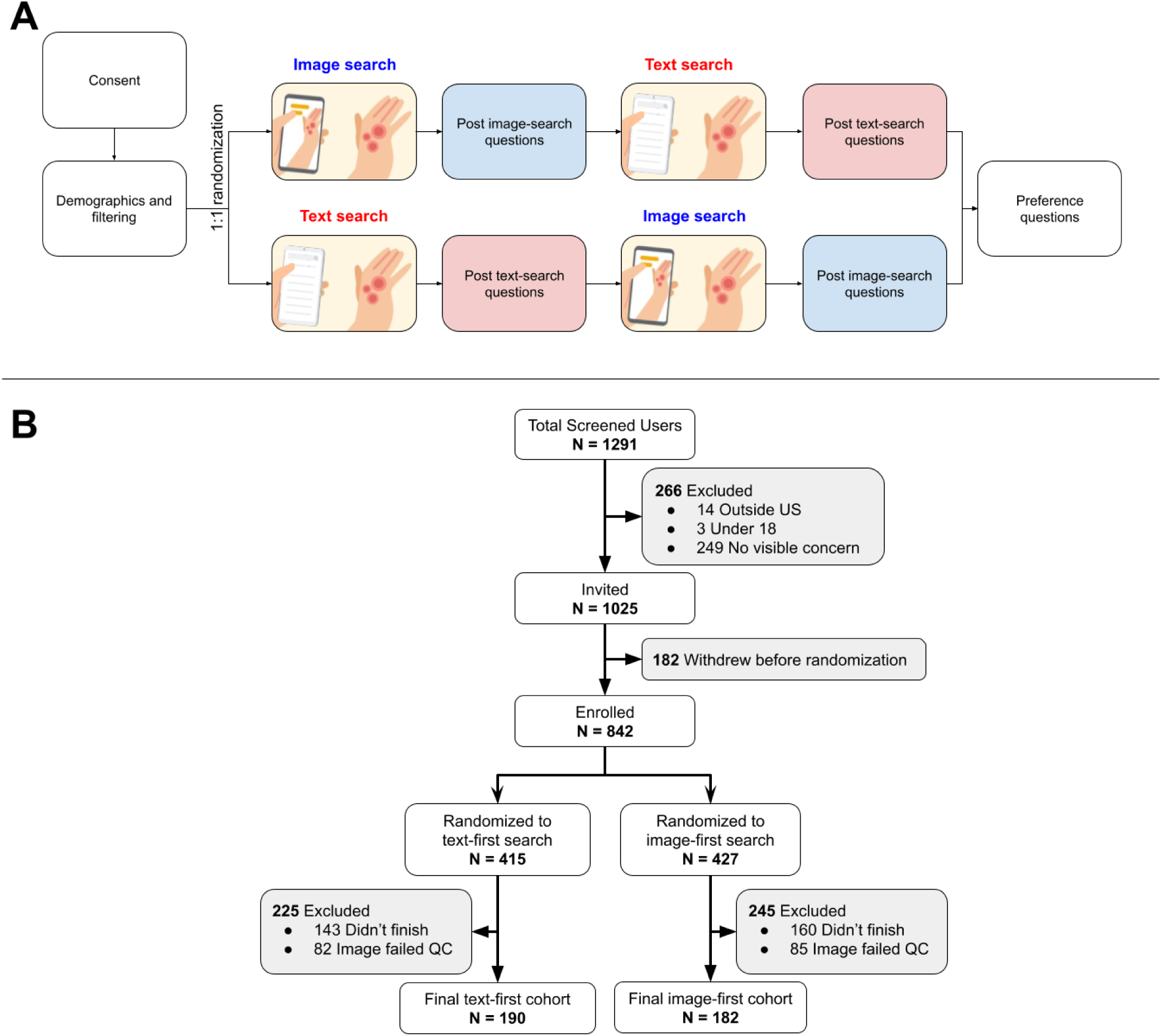
Study overview. **A** - App and survey workflow. Individuals with visible skin conditions were consented and enrolled from the Qualtrics panel and then randomized into either text-based or image-based search first, afterwards completing survey questions relating to their satisfaction with this experience. They then performed their search with the other modality, afterwards completing questions relating to satisfaction from this second experience, and comparing their preferences between the two experiences. **B** - STARD diagram depicting patient flow through the study.

Post randomization (to either text-based search or image-based search), respondents completed the first search, and were asked to rate their satisfaction along six different dimensions using a 5-point Likert scale ranging from “very dissatisfied” to “very satisfied”: (1) overall experience, (2) helpfulness of results, (3) ease of search, (4) ease of app use, (5) time taken to get any results, and (6) time taken to get helpful results. Respondents were then asked to conduct the second search of the same concern using the other experience (ie, those randomized to text-based search then completed image-based search and vice versa) and were asked to rate their satisfaction with the second experience along the same six dimensions using the same Likert scale. Finally respondents were asked whether they preferred the image-based or text-based search experiences for each of the same six dimensions. They were then asked what they would prefer to use for a future search: text-based search, image-based search, or both (including image-first or text-first) along several axes. Finally, an optional free text field was available for comments about the survey or feature.

### Statistical analyses

Statistical analyses were conducted using Python 3 and the Numpy, Pandas, and Statsmodels packages. Statistical significance was defined as p < 0.05.

All statistical tests were conducted using 2-sided permutation tests with 1000 iterations; all 95% confidence intervals (CIs) were based on bootstrapping with 1000 iterations. Items evaluated include: percentage of those at least somewhat satisfied with a given search experience in each of the six dimensions, percentages of those who preferred image-based vs text-based search in each of the six dimensions, and the percentage of those who preferred to use image-only, text-only, and both experiences in the future.

Additionally, separate multivariable logistic analysis were conducted to further analyze factors associated with: (1) satisfaction with image-based search, (2) those who prefer image-based search to text-based search, (3) those who wish to include image-based search in the future. When more than 2 subgroups were being compared, the subgroup with the largest number of respondents was chosen as the reference group.

### App details

Prior to completion of the survey, respondents were asked to install or update the publicly-available version of the Google app during the study period (October - December 2023) on their smartphone from the iOS app or Android Play stores. However, app versions were not verified in order to minimize friction for respondents. This app contains methods both for image-based search (Google Lens) and text-based search (Google Search). Google Lens takes as input a photo captured by the user and then displays a sliding “carousel” below of similar appearing skin conditions. Users can click on these to see results related to each one. Google Search takes a text query as input and returns a list of related links. See Figure 2 for screenshots showing the Google app using both Lens and Search.

**Figure 2:**
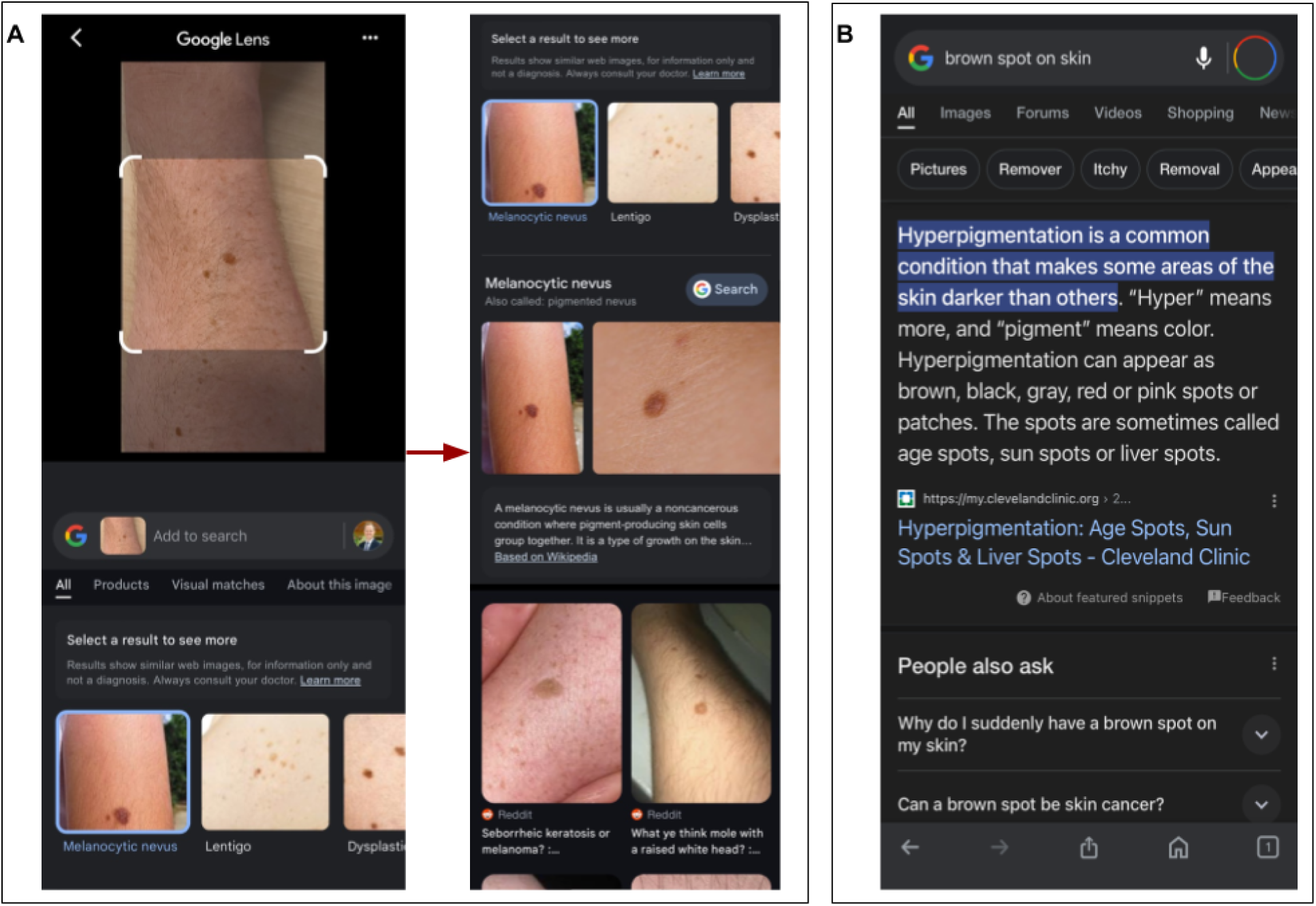
Screenshots of the Google app using A) image-based search (Google Lens) and B) text-based search (Google Search). Google Lens (**A**) takes as input a photo captured by the user and then displays a sliding “carousel” below of similar appearing skin conditions. Users can click on these to see a summary and results related to each one. Google Search (**B**) takes a text query as input and returns a list of related links, sometimes displaying relevant content from top results directly on the search page as shown in the screenshot above.

## Results

### Survey respondents

Demographics of survey respondents are shown in Table 1, including age brackets, gender, ethnicity, confidence in using the mobile app, education level, self-reported Fitzpatrick skin type (FST), duration of the concern, percentage of those who have not received a previous diagnosis for this concern, how much time had been spent searching this concern, and familiarity with text-based and image-based search. Percentages were similar between the randomized groups. Respondents spent a median of 11.2 minutes (interquartile range 8.0 - 16.2 minutes) completing the searches and survey.

**Table 1:** Demographics of respondents.

|  |  | All respondents | Image first study arm | Text first study arm |
| --- | --- | --- | --- | --- |
| Gender | Woman | 43.5% | 41.8% | 45.3% |
| Race | White | 85.8% | 85.9% | 85.7% |
| Ethnicity | Hispanic | 14.0% | 15.9% | 12.1% |
| Age | 18 to 24 | 27.7% | 24.2% | 31.1% |
|  | 25 to 34 | 17.5% | 17.0% | 17.9% |
|  | 35 to 44 | 14.0% | 17.0% | 11.1% |
|  | 45 to 54 | 15.3% | 16.5% | 14.2% |
|  | 55 to 64 | 12.9% | 15.9% | 10.0% |
|  | 65 or over | 12.6% | 9.3% | 15.8% |
| Tech literacy | Mobile app use confidence: extremely or very confident | 96.0% | 96.2% | 95.8% |
| Education level | Education level (at least Bachelors degree) | 61.6% | 61.0% | 62.1% |
| Fitzpatrick skin type | 1 | 11.8% | 12.1% | 11.6% |
|  | 2 | 37.4% | 36.3% | 38.4% |
|  | 3 | 33.9% | 35.7% | 32.1% |
|  | 4 | 8.6% | 8.8% | 8.4% |
|  | 5 | 6.2% | 6.0% | 6.3% |
|  | 6 | 1.6% | 0.5% | 2.6% |
|  | Other | 0.5% | 0.5% | 0.5% |
| Duration of concern | New concern | 30.6% | 31.9% | 29.5% |
|  | Recurring concern | 29.0% | 27.5% | 30.5% |
|  | Recent change | 17.7% | 18.1% | 17.4% |
|  | Long-time concern | 22.6% | 22.5% | 22.6% |
| Concern diagnosis status | No previous diagnosis | 22.6% | 20.3% | 24.7% |
| Time already spent | < 1 hour | 20.7% | 20.3% | 21.1% |
|  | Hours | 26.1% | 30.2% | 22.1% |
|  | < 30 minutes | 20.4% | 19.2% | 21.6% |
|  | Many days | 22.0% | 17.6% | 26.3% |
|  | None or a few seconds | 10.8% | 12.6% | 8.9% |
| Familiarity with image-based search | At least moderately familiar with image-based search | 73.1% | 70.3% | 75.8% |
|  | At least moderately familiar with derm image-based search | 63.5% | 62.6% | 64.2% |
| Familiarity with text-based search | At least moderately familiar with text-based search | 89.5% | 88.5% | 90.5% |
|  | At least moderately familiar with derm text-based search | 81.5% | 83.5% | 79.7% |

### Satisfaction with image-based and text-based search

The percentage of respondents at least somewhat satisfied with text-based and image-based search is shown in Table 2 and Figure 3. In general, a large majority of respondents were at least moderately satisfied with both experiences, with greater than 90% of respondents reporting this in each of the six dimensions. There were no statistically significant differences in satisfaction rates in any of the dimensions. Multivariable analysis of factors associated with satisfaction for image-based search (Supplemental Figure S1) showed women to be associated with decreased satisfaction (91.6% vs 97.0% satisfied, log odds −1.3 (95% CI −2.4, −0.19), p=0.022), and FST 3 or 4 to be associated with increased satisfaction relative to FST 1 or 2 (98.7% vs 90.7% satisfied, log odds 2.0 (95% CI 0.44, 3.6), p=0.012). There was no statistically significant difference for FST 5 or 6 vs 1 or 2.

**Figure 3:**
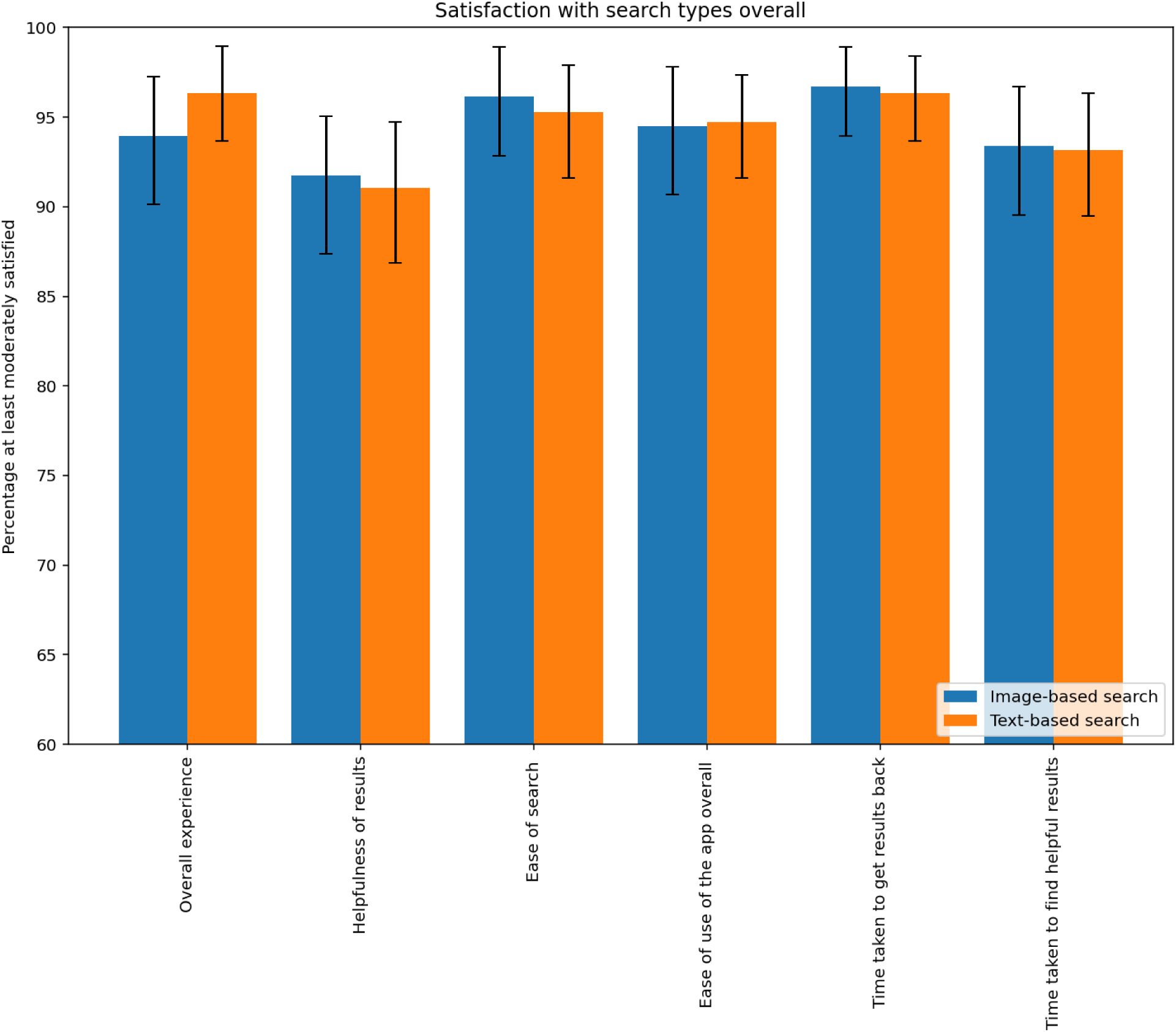
Percentage of respondents at least somewhat satisfied with Image or Text-based search in the following categories, only including the experience they were randomized to use first. Respondents were highly satisfied with both the image-based and text-based search experiences. There were no statistically significant differences between them.

**Table 2:** Percentage of respondents at least somewhat satisfied with Image or Text-based search in the following categories, only including the experience they were randomized to use first. Differences are bolded if statistically significant (p-value < 0.05).

|  |  | All respondents |
| --- | --- | --- |
| Overall experience | Image | 94.0% (90.1 - 97.3) |
|  | Text | 96.3% (93.7 - 98.9) |
|  | Difference | -2.4% (p=0.221) |
| Helpfulness of results | Image | 91.8% (87.4 - 95.1) |
|  | Text | 91.1% (86.8 - 94.7) |
|  | Difference | 0.7% (p=0.709) |
| Ease of search | Image | 96.2% (92.9 - 98.9) |
|  | Text | 95.3% (91.6 - 97.9) |
|  | Difference | 0.9% (p=0.613) |
| Ease of app use | Image | 94.5% (90.7 - 97.8) |
|  | Text | 94.7% (91.6 - 97.4) |
|  | Difference | -0.2% (p=0.827) |
| Time to get results | Image | 96.7% (94.0 - 98.9) |
|  | Text | 96.3% (93.7 - 98.4) |
|  | Difference | 0.4% (p=0.787) |
| Time to get helpful results | Image | 93.4% (89.5 - 96.7) |
|  | Text | 93.2% (89.5 - 96.3) |
|  | Difference | 0.2% (p=0.823) |

Respondents were highly satisfied with both the image-based and text-based search experiences. There were no statistically significant differences between them.

### Preference between image-based and text-based search

The percentage of respondents preferring text-based vs image-based search is shown in Figure 4A, with more detailed data listed in Supplemental Table S1. Respondents preferred image-based search with statistical significance in all dimensions except time to get helpful results, where there was still a trend towards preferring image-based search. For overall experience, 40.9% (35.8 - 45.7) preferred image-based search and 30.9% (26.3 - 35.8) preferred text-based search (difference 10.0%, p-value 0.004). An ordering effect was seen, with these effects strongly driven by large preferences for image-based search in those randomized to use this first, though trends in preference for image search were still seen in those randomized to text-based search first (roughly half the sample size), and in two of six dimensions these trends reached statistical significance. Multivariable analysis of factors associated with preferring image-based search (Supplemental Figure S2) showed age ≥ 45 (64.5% vs 51.9%, log odds 1.1 (95% CI 0.42, 1.7), p=0.001) and FST 3 or 4 (64.6% vs 51.9%, log odds 0.61 (95% CI 0.025, 1.2), p=0.041) associated with increased preference for image-based search and individuals who identified as women were associated with decreased preference for image-based search (46.2% vs 65.5%, log odds −0.97 (95% CI −1.5, −0.39), p=0.001). There was no statistically significant difference for FST 5 or 6 vs 1 or 2.

**Figure 4:**
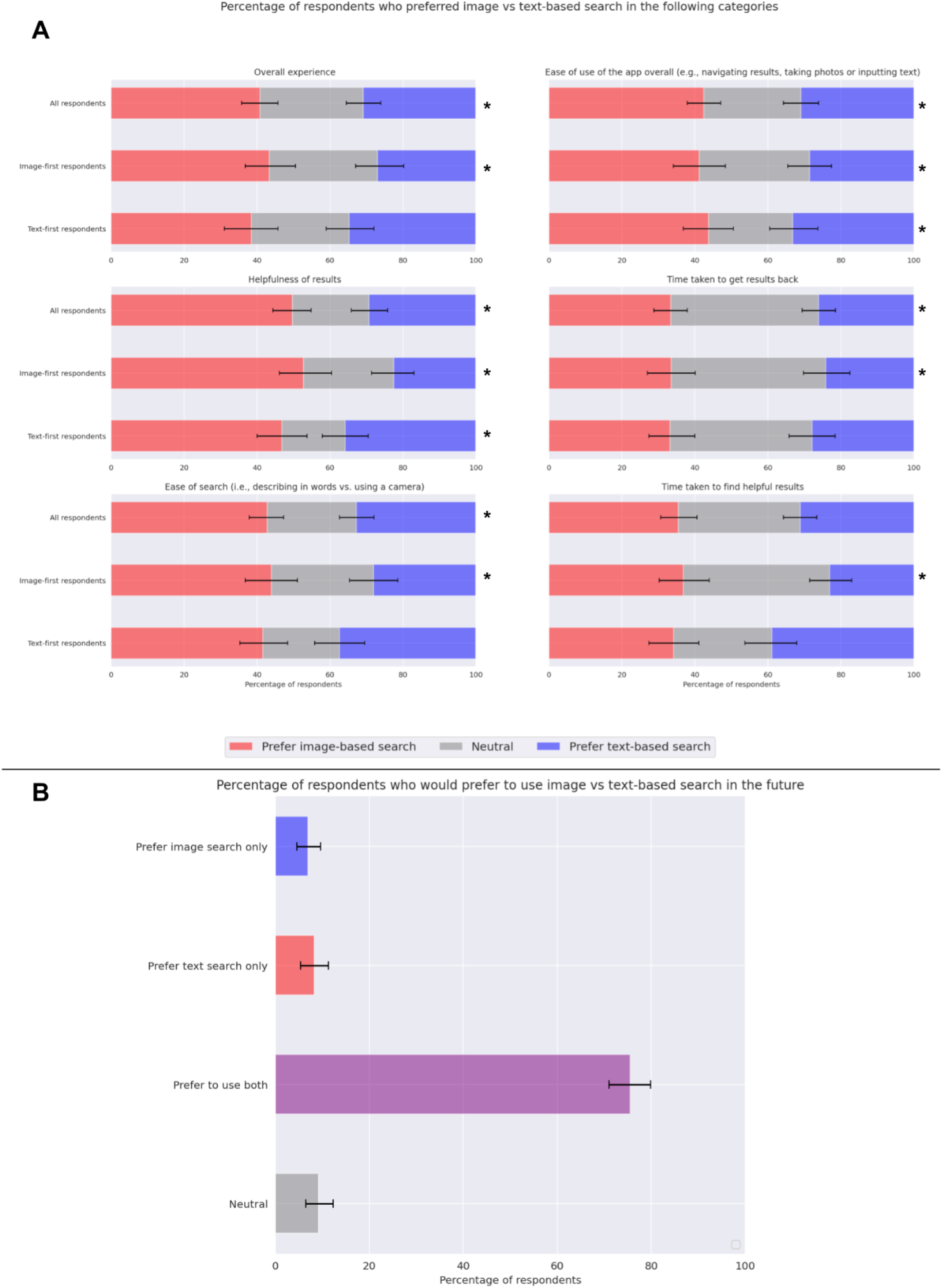
Respondents’ preferences for image-based vs text-based search. **A** - Percentage of respondents who preferred image-based vs text-based search in several categories. Statistically significant differences in preference are denoted by **\***. Respondents generally preferred image-based search to text-based search, but an ordering effect was observed, with this preference stronger for the respondents who used image-based search first. Of note, however, text-based search first respondents also preferred image-based first for “Helpfulness” and “Ease of App Use.” See Supplemental Table S1 for more detailed results. **B** - Preference for using image vs text-based search for a future search. The vast majority of respondents want to to include image search in the future, whether alone or in combination with text-based search. This clarifies the forced choice (image vs text) observations: respondents saw value in using both image and text search in combination.

### Future preferences

The percentage of respondents who would prefer including image-based, text-based, or both searches in the future is shown in Table 3 and Figure 4B. A large majority of respondents want to include image-based search in the future (82.5% (95% CI 78.2, 86.3)), with the majority of these preferring to include text-based search as well. Only 8.3% (95% CI 5.4, 11.3) preferred to use text-based search alone. Multivariable analysis of factors associated with preferring to include image-based search in the future, whether alone or in combination with text-based search, is shown in Supplemental Figure S3. Age ≥ 45 (95.1% vs 87.9%, log odds 1.8 (95% CI 0.75, 2.9), p<0.001), FST 3 or 4 (95.1% vs 87.4%, log odds 1.0 (95% CI 0.075, 2.0), p=0.035), and previous familiarity with image-based search (92.9% vs 86.3%, log odds 1.3 (95% CI 0.38, 2.3), p=0.006) were all associated with increased preference for including image-based search.

**Table 3:** Percentage of respondents who would prefer Image vs Text-based search in the future.

|  | **All respondents** | **Image first** | **Text first** |
| --- | --- | --- | --- |
| Neutral | 9.1% (6.5 - 12.4) | 7.7% (3.8 - 11.5) | 10.5% (6.3 - 15.3) |
| Prefer image only | 7.0% (4.6 - 9.7) | 8.2% (4.4 - 12.6) | 5.8% (2.6 - 8.9) |
| Prefer text only | 8.3% (5.4 - 11.3) | 6.0% (2.7 - 10.4) | 10.5% (6.3 - 14.7) |
| Prefer using both, image first | 48.4% (43.0 - 53.2) | 57.1% (50.5 - 64.3) | 40.0% (33.2 - 46.8) |
| Prefer using both, text first | 27.2% (22.8 - 32.0) | 20.9% (15.4 - 26.4) | 33.2% (26.3 - 39.0) |
| **Prefer using both overall** | 75.5% (71.0 - 79.8) | 78.0% (72.5 - 83.5) | 73.2% (66.8 - 79.5) |
| **Prefer including image search, either alone or with text search** | 82.5% (78.2 - 86.3) | 86.3% (81.3 - 91.2) | 78.9% (73.2 - 84.7) |

The majority of respondents, regardless of what search tool they used first, reported a preference for including image-based search in the future.

### Qualitative comments

Respondents’ free text comments covered several aspects. Many had a positive reaction to using image-based search: “I had no idea this was available in the google app, but now that i know i will certainly use it in the future!” and “I learned something new today and will share with friends. I have never used this and I liked it a lot! I can see where this could save me time in the future.” or felt image-based search to be an easier input modality than words: “I love the Google Lens search feature. It helps find things you might not be able to properly describe. I find it very helpful.” and “Google Lens has efficient search, more so when I don’t know the words to use in searching”. Others appreciated that the output modality of image-based search was images, helping them to understand search results: “I prefer the image option because it’s … easier to understand by seeing images of what you are searching”.

Reasons given for preferring to use both modalities included additional confidence: “The combination of image and text gives me more confidence in the research.”, and the benefits of greater precision using text: “I find it easier to search more precisely with text compared to with a picture.” Finally, desired areas of improvement included feature requests such as using the front-facing camera, improving focus quality for close-ups, and increasing the diversity of result images shown.

## Discussion

In this study we explored the experiences and attitudes of users interacting with image-based vs text-based search for skin concerns. We found that generally respondents were highly satisfied with each experience, but that they preferred image-based search with regards to helpfulness, ease of search, ease of app use, time to get results, and the overall experience. We also found that respondents overwhelmingly (>75%) wished to include image-based search in their future queries, generally in combination with text-based search as well. This helps explain the trends observed in the forced-choice preference questions (image vs. text) were the differences were smaller; respondents saw value in using both modalities in combination.

This work builds on previous work, both within and outside of our institution, into the automated detection and differentiation of skin conditions from photos via artificial intelligence. In 2017, Esteva et al. reported dermatologist-level classification of skin conditions with a convolutional neural network-based model utilizing a dataset of more than 100,000 photos representing more than 2,000 different skin conditions^10^. This result has subsequently been replicated and expounded upon many times^11–17^, demonstrating that reliable detection and classification of skin conditions is possible with AI-based methods.

Significantly, many AI-based dermatology applications in the past have been shown to be subject to concerning amounts of bias, especially in regards to those with darker skin tones^18–21^. Importantly in this work we demonstrate that respondents with darker skin types are at least as satisfied with the image-based search experience as those with FST 1 or 2, and find no differences in satisfaction or preferences between those who self-identify as non-Hispanic White versus others.

Our study’s randomized setup also revealed an important aspect of ordering in this experiment with two search experiences per respondent. Specifically, while respondents on average preferred using both image- and text-based search for future concerns, with most of these being using image search prior to text search, this effect was stronger for respondents who were randomized to using image based search first. This could reflect a familiarity or anchoring effect. As such, a sequential study design (where all respondents first use the traditional text-based search, followed by the less-familiar image-based search) may not be appropriate, by virtue of skewing the observed effect size.

This work has several limitations. Firstly, as this was a user experience study to study user satisfaction, preference and sentiments, we did not capture personal health information or obtain respondents’ photos of their conditions and therefore, the “accuracy” of the results they received could not be verified. This was an intentional choice as this product is designed to be used for information gathering, similar to text-based search, and is not meant to provide diagnoses. Similarly, the inability to obtain an accurate “ground truth” would have hindered attempts to judge the appropriateness of self-reported next steps. Thus future work will be needed to examine in-depth if, how, and why users change decisions based on online information. Additionally, the survey respondents’ concerns represented a mix of those who had already been diagnosed by a physician and those who had new concerns (Table 1), and as this mixture may not be representative of the distribution of real users, may influence the results. In addition, as the respondents here were recruited via an online-based platform (Qualtrics), it is possible that these users may be more tech-savvy or willing to adopt new technology than the average real world user. Finally, as compared to survey respondents, real-world users who have an urgent concern may have different concerns and priorities at baseline, leading to possible differences in reported satisfaction.

In summary, this work demonstrates that users across demographics included are highly satisfied with an AI-based tool allowing image-based searching for visible skin conditions. Such a tool holds promise in helping users gather information about their skin conditions so that they can be more well-informed regarding their dermatological health.

## Data Availability

Data produced in the present work may be made available upon reasonable request to the authors.

## Supplemental Tables

**Supplemental Table S1:** Percentage of respondents who preferred Image vs Text-based search in the following categories. Differences are bolded if statistically significant (p < 0.05).

|  |  | All respondents | Image first | Text first |
| --- | --- | --- | --- | --- |
| Overall experience | Prefer image | 40.9% (35.8 - 45.7) | 43.4% (36.8 - 50.5) | 38.4% (31.1 - 45.8) |
|  | Prefer text | 30.9% (26.3 - 35.8) | 26.9% (20.9 - 34.1) | 34.7% (28.4 - 41.6) |
|  | Neutral | 28.2% (24.2 - 32.8) | 29.7% (23.6 - 36.3) | 26.8% (20.5 - 33.7) |
|  | Difference* | **9.9% (p=0.004)** | **16.5% (p=0.002)** | 3.7% (p=0.405) |
| Helpfulness | Prefer image | 49.7% (44.4 - 54.8) | 52.7% (46.1 - 60.4) | 46.8% (40.0 - 53.7) |
|  | Prefer text | 29.3% (24.5 - 34.4) | 22.5% (16.5 - 28.0) | 35.8% (29.5 - 42.1) |
|  | Neutral | 21.0% (16.9 - 25.0) | 24.7% (18.7 - 31.3) | 17.4% (12.1 - 22.6) |
|  | Difference* | **20.4% (p<0.001)** | **30.2% (p<0.001)** | **11.1% (p=0.031)** |
| Ease of Search | Prefer image | 42.7% (37.9 - 47.3) | 44.0% (36.8 - 51.1) | 41.6% (35.3 - 48.4) |
|  | Prefer text | 32.8% (28.2 - 37.6) | 28.0% (21.4 - 34.6) | 37.4% (30.5 - 44.2) |
|  | Neutral | 24.5% (19.9 - 28.8) | 28.0% (22.0 - 35.2) | 21.1% (15.8 - 27.4) |
|  | Difference* | **9.9% (p=0.002)** | **15.9% (p<0.001)** | 4.2% (p=0.342) |
| Ease of App Use | Prefer image | 42.5% (37.9 - 47.0) | 41.2% (34.1 - 48.4) | 43.7% (36.8 - 50.5) |
|  | Prefer text | 30.9% (26.1 - 35.8) | 28.6% (22.5 - 34.6) | 33.2% (26.8 - 40.0) |
|  | Neutral | 26.6% (22.3 - 30.9) | 30.2% (23.1 - 36.8) | 23.2% (16.8 - 28.9) |
|  | Difference* | **11.6% (p<0.001)** | **12.6% (p=0.011)** | **10.5% (p=0.029)** |
| Time for results | Prefer image | 33.3% (28.8 - 37.9) | 33.5% (26.9 - 40.1) | 33.2% (27.4 - 40.0) |
|  | Prefer text | 26.1% (21.5 - 30.6) | 24.2% (18.1 - 30.8) | 27.9% (21.6 - 34.2) |
|  | Neutral | 40.6% (36.0 - 45.7) | 42.3% (34.6 - 50.0) | 38.9% (31.6 - 45.8) |
|  | Difference* | **7.3% (p=0.023)** | **9.3% (p=0.035)** | 5.3% (p=0.224) |
| Time for helpful results | Prefer image | 35.5% (30.6 - 40.6) | 36.8% (30.2 - 44.0) | 34.2% (27.4 - 41.1) |
|  | Prefer text | 31.2% (26.6 - 35.8) | 23.1% (17.6 - 29.1) | 38.9% (31.6 - 45.8) |
|  | Neutral | 33.3% (28.5 - 38.2) | 40.1% (33.0 - 47.8) | 26.8% (21.1 - 33.2) |
|  | Difference* | 4.3% (p=0.169) | **13.7% (p=0.001)** | -4.7% (p=0.280) |

Respondents generally preferred image-based search to text-based search. An ordering effect was observed, with this preference most highly associated with those who used image-based search first, although of note text-based search first respondents also preferred image-based first for “Helpfulness” and “Ease of App Use.” \*Difference indicates Prefer image - Prefer text.

## Supplemental Figures

**Supplemental Figure S1:**
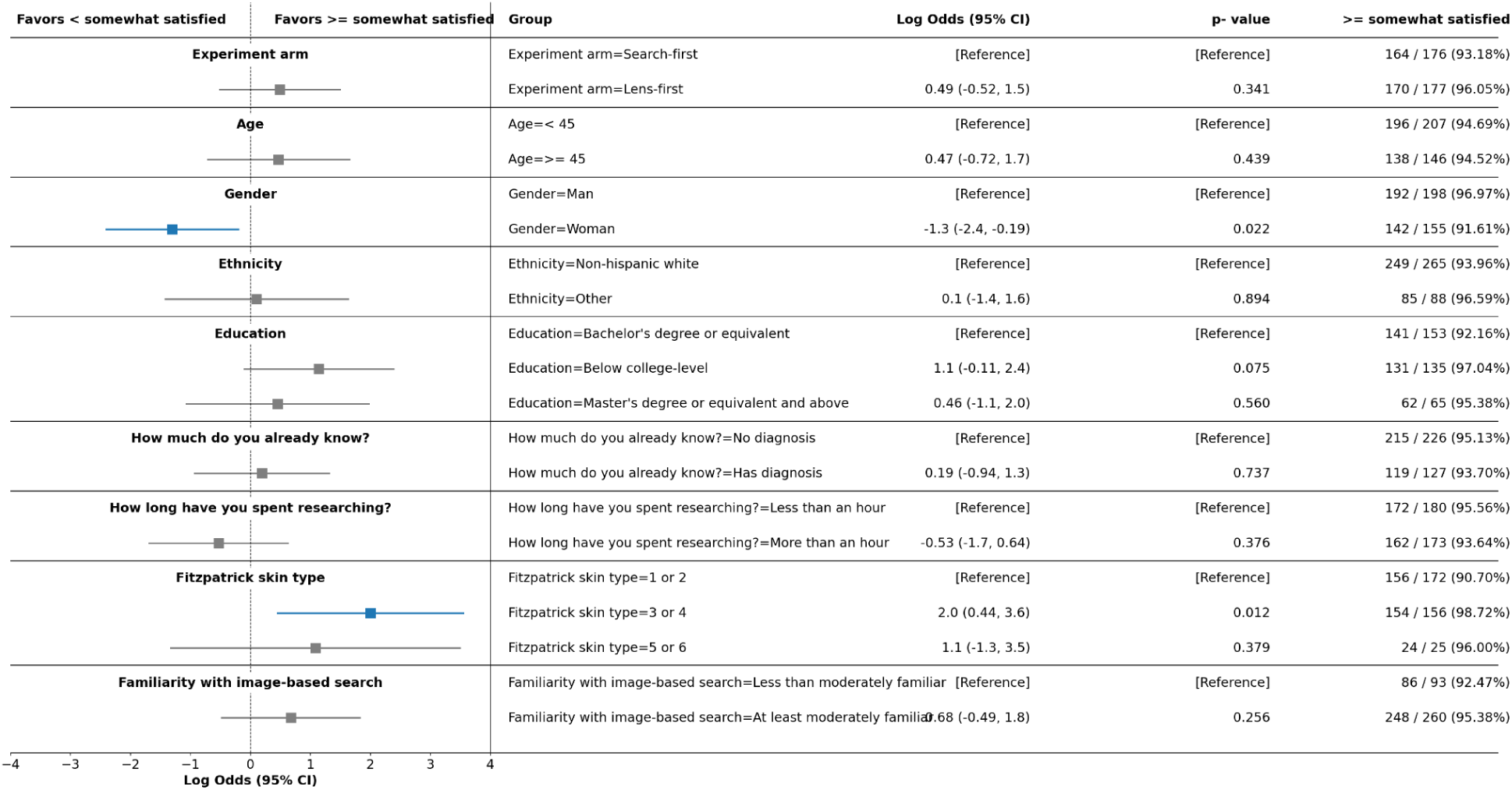
Multivariable logistic regression analysis predicting respondents at least “somewhat satisfied” with image-based search. All groups were overwhelmingly satisfied, but respondents who identified as women were less likely to be satisfied vs those who identified as men. Additionally, respondents with Fitzpatrick skin types 3 or 4 were more likely to be satisfied compared to those with types 1 or 2.

**Supplemental Figure S2:**
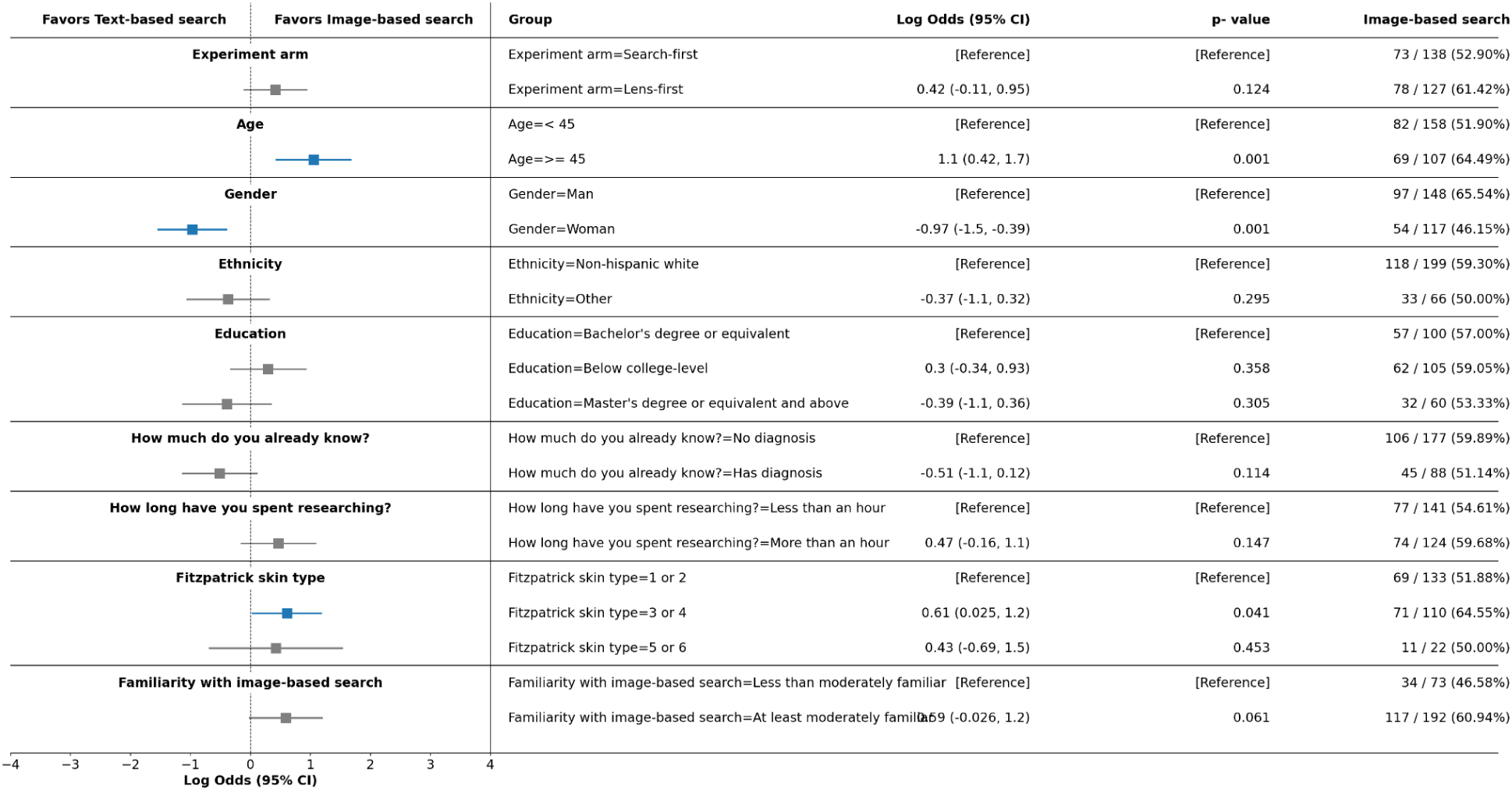
Multivariable logistic regression analysis predicting respondents who preferred image vs text-based search overall. Patients older than 45 and those with Fitzpatrick skin types 3 or 4 were more likely to prefer image-based search. Respondents who identified as women were less likely to prefer image-based search.

**Supplemental Figure S3:**
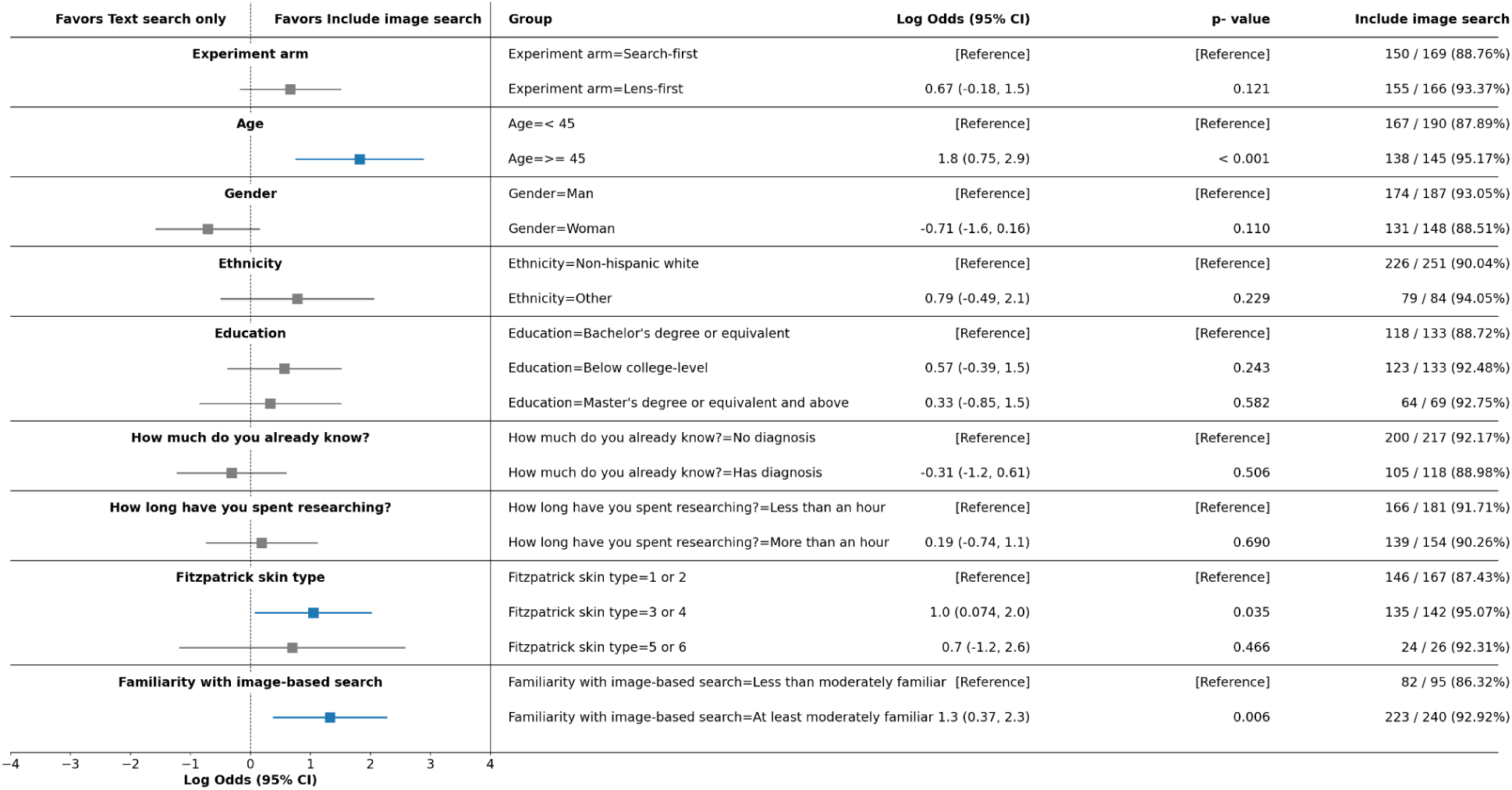
Multivariable logistic regression analysis predicting respondents who prefer to include image-based search in future searches (either alone or with text-based search) vs those who would prefer text-based search only. A large majority of all groups favored including image-based search in the future. Respondents older than 45, those with Fitzpatrick skin types 3 or 4, and those who were at least moderately familiar with image-based search at the start of the study were more likely to want to include image-based search in the future.

